# Protocol for a seamless phase 2A-phase 2B randomized double-blind placebo-controlled trial to evaluate the safety and efficacy of benfotiamine in patients with early Alzheimer’s disease (BenfoTeam)

**DOI:** 10.1101/2024.04.19.24306070

**Authors:** Howard H. Feldman, José A. Luchsinger, Gabriel C. Léger, Curtis Taylor, Diane M. Jacobs, David P. Salmon, Steven D. Edland, Karen Messer, Carolyn Revta, Sarah A. Flowers, Kerry S. Jones, Albert Koulman, Kevin E. Yarasheski, Philip B. Verghese, Venky Venkatesh, Henrik Zetterberg, January Durant, Jody-Lynn Lupo, Gary E. Gibson, the ADCS BenfoTeam Study Group

## Abstract

**Background:** Benfotiamine provides an important novel therapeutic direction in Alzheimer’s disease (AD) with possible additive or synergistic effects to amyloid targeting therapeutic approaches.

**Objective:** To conduct a seamless phase 2A-2B proof of concept trial investigating tolerability, safety, and efficacy of benfotiamine, a prodrug of thiamine, as a first-in-class small molecule oral treatment for early AD.

**Methods:** This is the protocol for a randomized, double-blind, placebo-controlled 72-week clinical trial of benfotiamine in 406 participants with early AD. Phase 2A determines the highest safe and well-tolerated dose of benfotiamine to be carried forward to phase 2B. During phase 2A, real-time monitoring of pre-defined safety stopping criteria in the first approximately 150 enrollees will help determine which dose (600 mg or 1200 mg) will be carried forward into phase 2B. The phase 2A primary analysis will test whether the rate of tolerability events (TEs) is unacceptably high in the high-dose arm compared to placebo. The primary safety endpoint in phase 2A is the rate of TEs compared between active and placebo arms, at each dose. The completion of phase 2A will seamlessly transition to phase 2B without pausing or stopping the trial. Phase 2B will assess efficacy and longer-term safety of benfotiamine in a larger group of participants through 72 weeks of treatment, at the selected dose. The co-primary efficacy endpoints in phase 2B are CDR-Sum of Boxes and ADAS-Cog13. Secondary endpoints include safety and tolerability measures; pharmacokinetic measures of thiamine and its esters, erythrocyte transketolase activity as blood markers of efficacy of drug delivery; ADCS-ADL-MCI; and MoCA.

**Conclusion:** The BenfoTeam trial utilizes an innovative seamless phase 2A-2B design to achieve proof of concept. It includes an adaptive dose decision rule, thus optimizing exposure to the highest and best-tolerated dose.

**Trial Registration:** ClinicalTrials.gov identifier: NCT06223360, registered on January 25, 2024.

## Introduction

Alzheimer’s disease (AD) is a progressive age-associated neurodegenerative disease. It is the most common cause of dementia and has devastating socioeconomic implications. More than 5 million people are afflicted with AD and related dementias in the United States, and by 2050 this number could rise as high as 16 million, with annual costs of over $1 trillion [1]. The potential to preserve, or even improve, cognition in adults at high risk of cognitive decline due to AD clearly has important implications, not only for the affected individual, but also for the support system that bears the social and financial burdens of long-term caregiving. Despite recent advances in amyloid targeting therapies, they do not address processes that are upstream and downstream of the amyloid cascade. Thus, complementary therapies that address these processes are needed.

Benfotiamine a prodrug of thiamine provides a novel therapeutic direction in AD that has potential for additive or synergistic effects beyond current mainstream approaches. It is a first-in-class small molecule with a unique mechanism of action, that raises blood thiamine (vitamin B1) 50-100 times to pharmacological levels. In this way, it addresses and treats a well-characterized tissue thiamine action deficiency in AD that is associated with changes in glucose metabolism and thiamine dependent post-translational modifications that are linked to AD pathology. This includes action on the α-Ketoglutarate Dehydrogenase Complex (KGDHC)-regulated succinylation of the TCA cycle that links brain glucose metabolism to AD pathophysiology [2]. These related post translational modifications have been identified on tau and Abeta with effects that have been shown to promote plaque and tangle formation [3], neuroinflammation [4, 5], neurodegeneration [2], and increased advanced glycation end products (AGE) [6] which are regulated by thiamine. AGE are toxic modifications that form by the chemical addition of glucose and its byproducts to proteins, lipids and nucleotides when glucose is not tightly controlled in the cell, as is seen with the abnormal glucose metabolism in AD. High concentrations of AGE are predictive of long-term decline in cognition-related daily living performance in patients with AD [7]. Whereas thiamine deficiency increases AGE, increasing brain thiamine activates transketolase, which diminishes AGE [8, 9].

CNS thiamine deficiency and its effects on memory and cognition have been well known since 1930s [10]. It is the cause of the amnestic disorder of the Wernicke-Korsakoff syndrome [10] with data strongly supporting its pathogenetic role as well in AD. Thiamine dependent processes are deficient in autopsied brains from AD patients including transketolase (controls the pentose phosphate pathway), the pyruvate dehydrogenase complex [PDHC, links glycolysis to the tricarboxylic acid cycle (TCA)], glyoxalase (detoxification of AGE precursors) and KGDHC (controls the TCA cycle) [11, 12]. Furthermore, the reduced thiamine dependent enzyme activity measured in post-mortem brain tissue in AD patients, correlates with pre-mortem degree of impairment on the Clinical Dementia Rating (CDR) score [13]. Preclinical studies using a range of models including isolated proteins, cell systems and animal models demonstrate that thiamine deficiency stimulates many AD-like pathological changes including amyloid plaque and tangle formation, neuronal loss, inflammation, oxidative stress, cholinergic deficits, abnormally reduced glucose metabolism and memory deficits [14].

Recent data provides a mechanistic framework for the decline in thiamine-dependent processes in the brains of patients with AD. Thiamine entry into the brain is controlled by transporters and the transporters are diminished in autopsied brains from AD patients and in brains from mouse models of AD [15]. High concentrations of thiamine in the serum such as those induced by benfotiamine can overcome a reduction in transporters [16]. Model systems studies have demonstrated that high thiamine achieved by increasing dosages of benfotiamine reverses AD-like changes including restoring thiamine-dependent enzymes, with improvement in brain glucose metabolism, in reducing plaques [17] and tangles [3], neuroinflammation [4, 5] neurodegeneration [2], and reducing AGE [6].

These preclinical studies have supported a small, single-site 12-month pilot trial of benfotiamine in persons with early AD (35 per treatment arm) that demonstrated benfotiamine is safe and well tolerated with encouraging PK and PD responses [6]. The treatment delivery of benfotiamine in a dose of 600 mg/d achieved its intended PK outcome with a 161-fold mean increase in blood thiamine. The trial also provided preliminary evidence of efficacy of benfotiamine on cognitive and functional outcomes. While group difference in change on the Alzheimer’s Disease Assessment Scale-Cognitive Subscale (ADAS-Cog) did not reach statistical significance, there was a numerical benefit, with 43% less decline in benfotiamine (-1.39) vs placebo (-3.26) (mixed effect model p = 0.071, GEE p = 0.137, and non-parametric Wilcoxon rank sum test p = 0.098). The CDR, a composite of cognition and everyday function, showed a significant benefit (p=0.034), and other measures showed favorable treatment effects on patterns of brain glucose utilization (p=0.002) and patterning scores (p=0.0329) on ^18^F- fluorodeoxyglucose positron-emission tomography (FDG PET). Plasma AGE, marking pharmacodynamic target engagement, were also reduced in a randomly selected subset of patients (p=0.044) [6]. In this study there was a pattern of pharmacogenomic response where participants without *APOE* ε4 were more responsive to benfotiamine than those with *APOE* ε4.

These preliminary results justify the testing of benfotiamine in a larger seamless phase 2A-phase 2B randomized controlled trial (RCT) to investigate its safety, tolerability, and efficacy in early AD [18]. The BenfoTeam trial will provide the next level of necessary clinical, safety and efficacy evidence to establish “Proof of Concept” (POC) for benfotiamine.

## Materials and methods

### Objectives

The primary objective of phase 2A is to determine the highest safe and well-tolerated daily dose of benfotiamine (600 mg/d or 1200 mg/d), as evaluated by the rate of tolerability events (TEs), to advance to the longer-term phase 2B study. The secondary objective of phase 2A is to evaluate additional measures of safety and tolerability of 600 mg/d or 1200 mg/d doses of benfotiamine.

The primary objective of phase 2B is to assess the efficacy of benfotiamine on global function and cognition over 72 weeks. The secondary objectives of phase 2B are to evaluate: 1) longer-term safety and tolerability of benfotiamine treatment, 2) the effects of benfotiamine’s PK relationships to primary clinical outcome measures, (plasma thiamine, and thiamine esters in whole blood, thiamine diphosphate (ThDP) and thiamine monophosphate (ThMP) and erythrocyte transketolase activity), and 3) benefits of benfotiamine on everyday activities of daily living and specific aspects of cognitive function (e.g., memory) over 72 weeks.

Exploratory objectives of phase 2B are to evaluate: 1) the downstream biological effects of treatment with benfotiamine on measures of neurodegeneration (cortical thickness MRI, plasma neurofilament light chain, total tau), neuroinflammation (plasma glial fibrillary acid protein) and AD pathophysiology (plasma p-tau231, p-tau217, Aβ 42/40, p-tau217/np-tau217(p- tau217 ratio) over 72 weeks, 2) the effects of benfotiamine on neuropsychiatric symptoms and cognitive-functional composite measures, 3) the effects of benfotiamine on remotely-administered measures of cognition, and to compare the sensitivity of remote and in-person cognitive assessments, and 4) the relationship between PK measures, including plasma thiamine, thiamine esters in whole blood (ThDP and ThMP), erythrocyte transketolase activity, and PD measures including AGE.

### Design

This overall design is a placebo controlled parallel group RCT over 72 weeks. The Phase 2A safety and dose selection decision will be reached when either 21 total TEs have been observed across both the high dose arm and the placebo arm, or when 160 person-months of exposure have accumulated in each of the high dose and placebo arms, whichever comes first (see S1 File). If the tolerability stopping rule is not reached by this predetermined number of TEs, the 1200 mg dose will continue, the 600 mg dose will be titrated upwards, and randomization will be adjusted for the remainder of trial enrollment to be 1:1 active versus placebo across the trial population. The completion of phase 2A will seamlessly transition to phase 2B without pausing or stopping the trial. Phase 2B will assess efficacy and longer-term safety of benfotiamine at the selected dose in a larger group of participants through 72 weeks of treatment.

The schedule of enrollment, interventions, and assessments is shown in Fig 1. The study schematic is shown in Fig 2.

**Figure 1.**
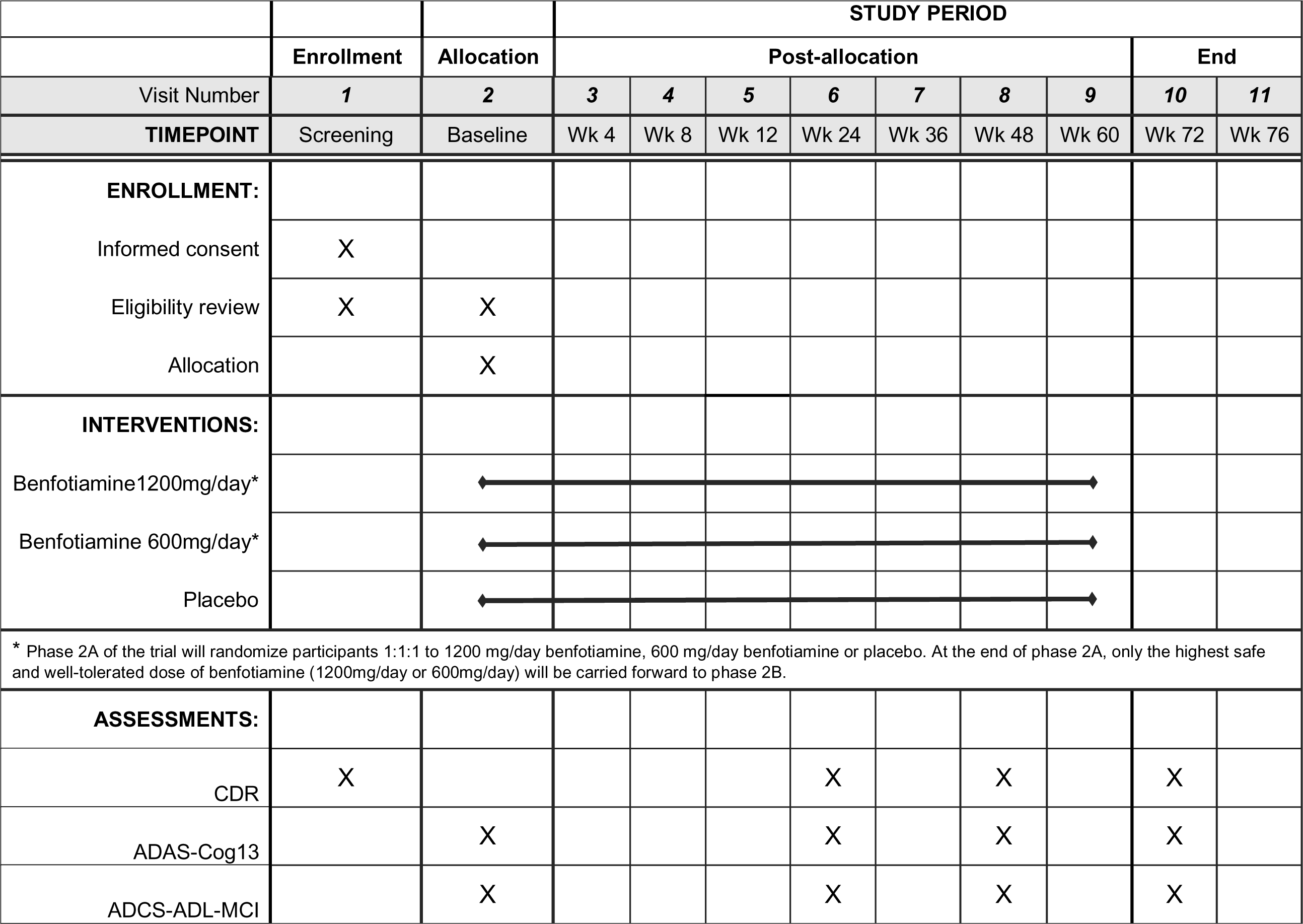
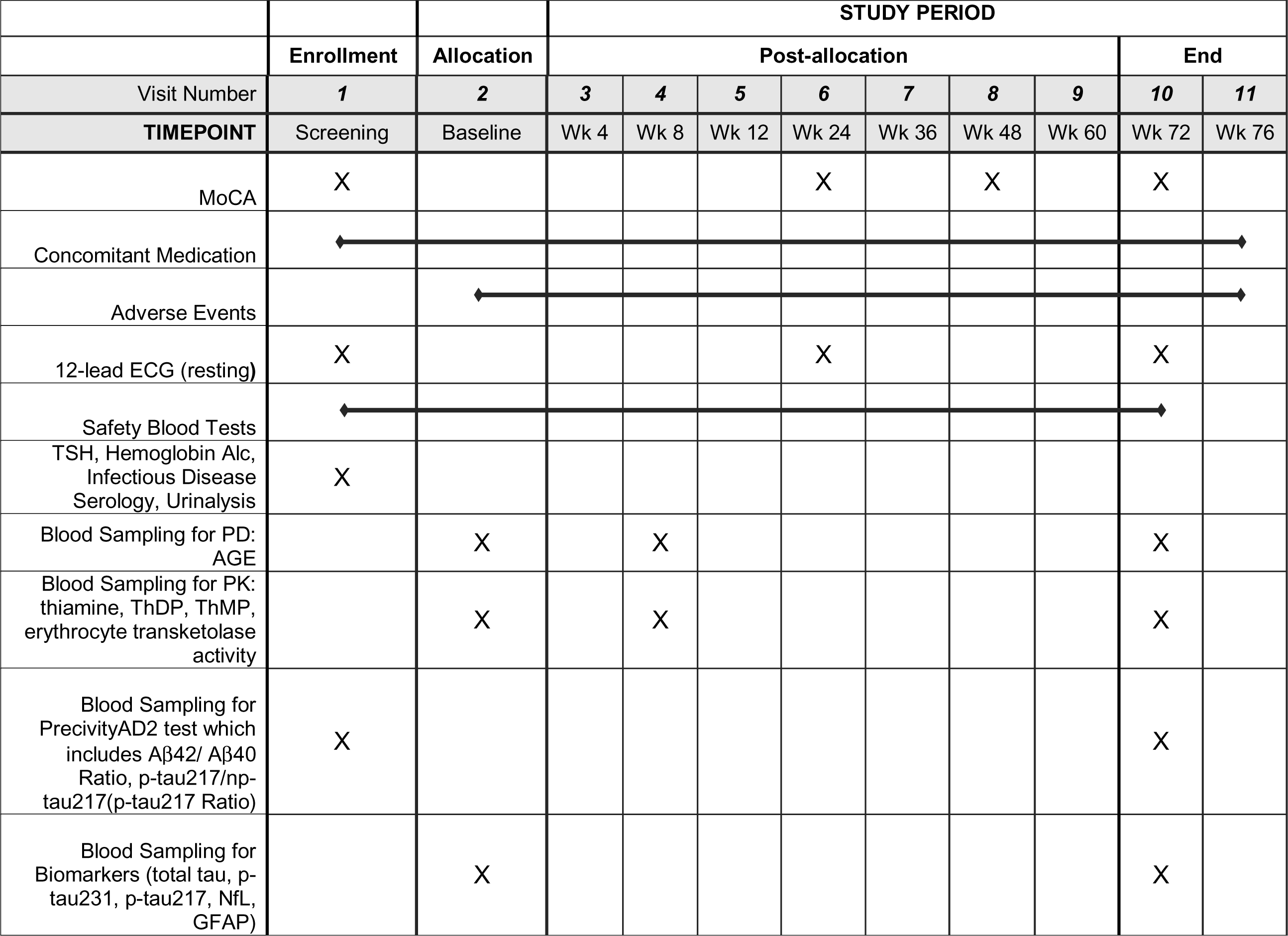
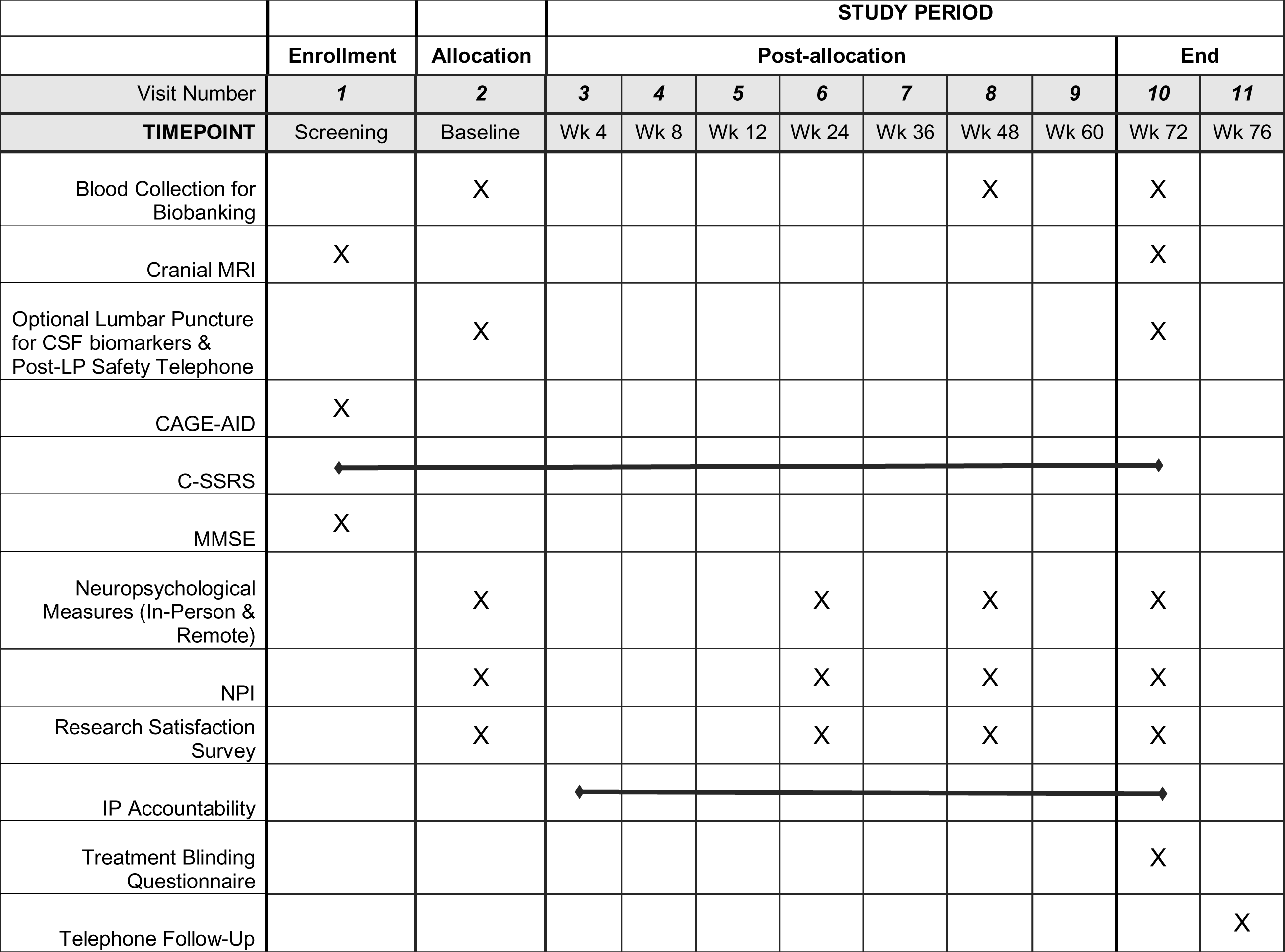
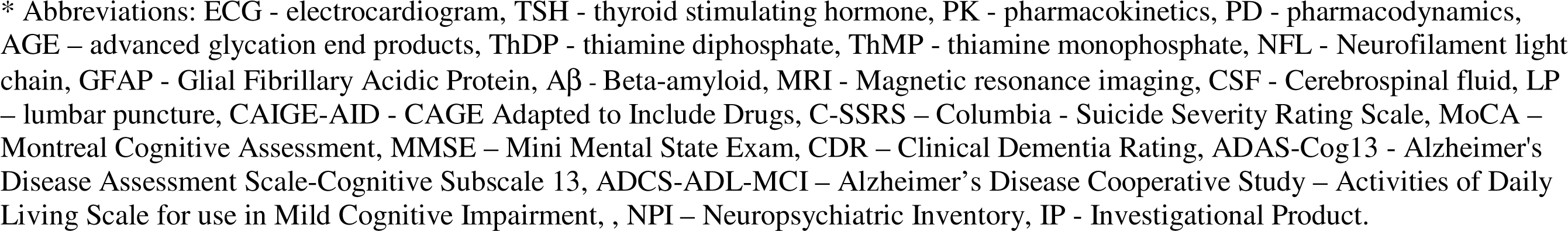
Schedule of enrollment, interventions, and assessments.

**Figure 2.**
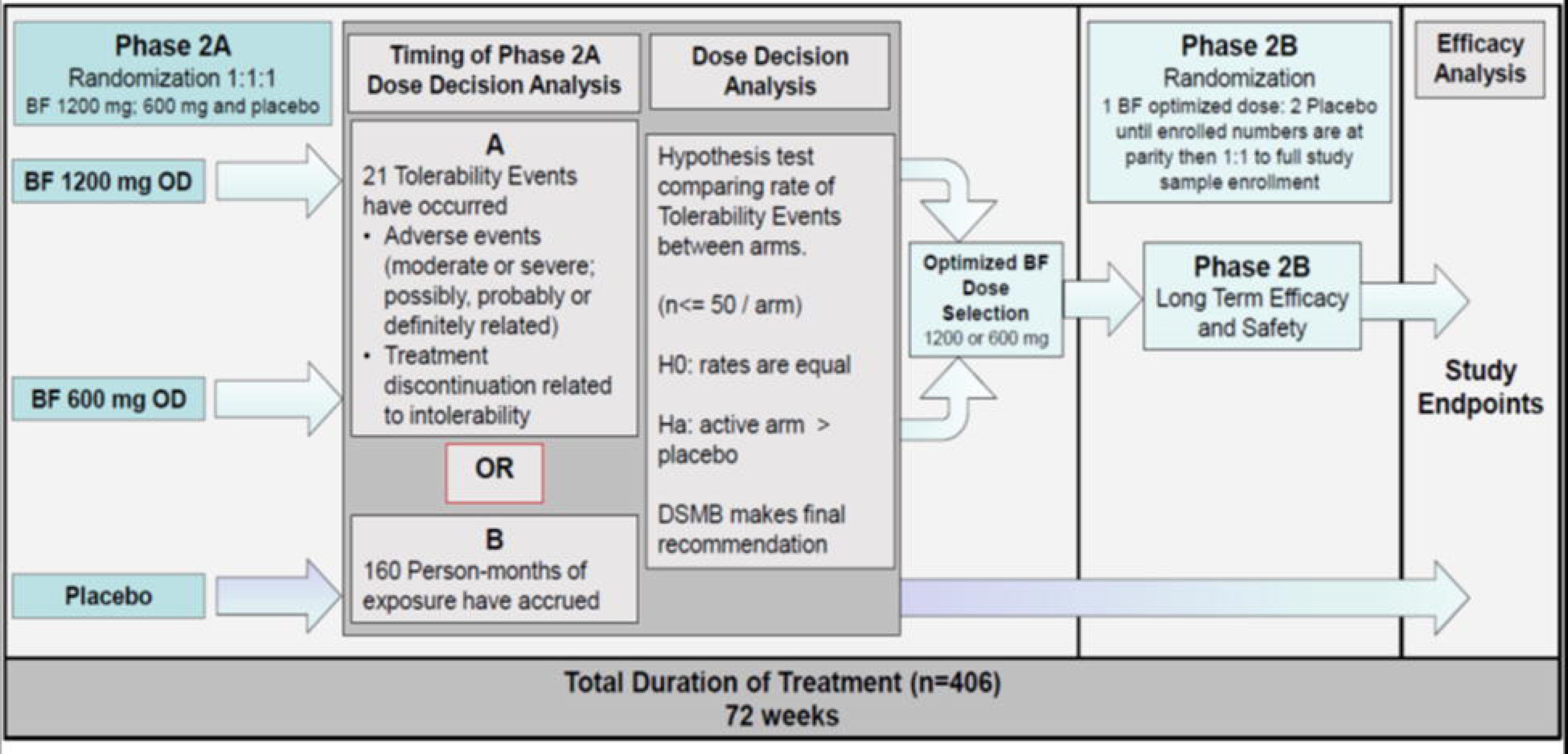
Study schematic.

### Participants

This trial (NCT06223360) will include 406 participants with early AD, including participants with MCI or mild dementia due to AD according to NIA-AA Diagnostic Criteria with a positive plasma AD biomarker test result (C_2_N PrecivityAD2; APS2 value >47.5) [19–22]. Key inclusion criteria include aged 50-89, Mini-Mental State Examination (MMSE) total score 20-30, Montreal Cognitive Assessment (MoCA) <26, and CDR score of 0.5 or 1 with memory score of ≥ 0.5 at screening. A complete list of inclusion criteria is presented in Table 1. Key exclusion criteria include any significant neurological or neurodegenerative disorder (other than AD), uncontrolled depression or major depression, a current diagnosis of uncontrolled type I or type II diabetes mellitus, a diagnosis of cancer (unless no evidence of recurrence for 5 years), any other major psychiatric disorder or active medical condition that impairs cognition or impacts participation, MRI at screening shows evidence of infection, tumor, cortical infarction, or multiple lacunes in prefrontal or critical memory regions, and initiation of a monoclonal antibody treatment targeting brain amyloid (including lecanemab, aducanumab, solanezumab, donanemab) within 6 months. A list of exclusion criteria is presented in Table 2. Recruitment and enrollment of participants will occur through the Alzheimer’s Disease Cooperative Study (ADCS) network at approximately 50 sites in the United States. A list of participating sites can be viewed at www.clinicaltrials.gov.

**Table 1.** Inclusion criteria.

| Inclusion criteria |  |
| --- | --- |
| 1. | Aged 50 to 89 (inclusive) at screening. |
| 2. | Mild Cognitive Impairment due to AD or Mild probable AD (NIA-AA<sup>1,2</sup>). |
| 3. | Mini-Mental State Examination (MMSE) score 20-30 inclusive at screening. |
| 4. | Montreal Cognitive Assessment score (MoCA) < 26 at screening. |
| 5. | Clinical Dementia Rating (CDR) global score of 0.5 or 1 with memory score of $\geq 0.5$ at screening. |
| 6. | Positive plasma AD biomarker signature:<br>Plasma test (C<sub>2</sub>N PrecivityAD2). To qualify participants will have a High Amyloid Probability Score (APS2 >47.5).<br>If previously treated with any anti-amyloid monoclonal antibody (AAMA) and C2N PrecivityAD2 is negative, documentation of a pre-AAMA treatment positive amyloid status with either amyloid PET, CSF, or appropriate plasma test. |
| 7. | Stable dosage of AChEIs and/or memantine for at least 3 months prior to screening. |
| 8. | Availability of a study partner who has frequent interaction with the participant (approximately >3-4 times per week) and is available for all study visits. |
| 9. | For FDA-approved monoclonal antibodies targeting brain amyloid (including lecanemab, aducanumab, solanezumab, donanemab), participants must be on stable dose of treatment for at least 6 months prior to screening. |
| 10. | Women must be post-menopausal for at least one year or surgically sterile (bilateral tubal ligation, hysterectomy, or bilateral oophorectomy) for at least 6 months prior to screening. |
| 11. | Fluent in English or Spanish. |
| 12. | Living in the community (includes assisted living facilities). |
| 13. | Ambulatory, or able to walk with an assistive device. |
| 14. | Informed consent from the participant (or the participant's legally authorized representative if lacking decisional capacity), as well as study partner consent. |

**Note.** <sup>1</sup> Albert, M.S., et al., The diagnosis of mild cognitive impairment due to Alzheimer's disease: recommendations from the National Institute on Aging-Alzheimer's Association workgroups on diagnostic guidelines for Alzheimer's disease. *Alzheimers Dement*, 2011. 7(3): p. 270-9. <sup>2</sup> McKhann, G.M., et al., The diagnosis of dementia due to Alzheimer's disease: Recommendations from the National Institute on Aging-Alzheimer's Association workgroups on diagnostic guidelines for Alzheimer's disease. *Alzheimer's & Dementia*, 2011. 7(3): p. 263-269.

**Table 2.** Exclusion criteria.

| Exclusion Criteria |  |
| --- | --- |
| 1. | Significant neurological disorder other than AD including hypoxia, stroke, traumatic brain injury. |
| 2. | Diagnosis of uncontrolled depression or major depression, as determined by DSM-IV, unless successfully treated. |
| 3. | Any other major psychiatric disorder that impairs cognition or impacts participation in the study is exclusionary. Participants with major psychiatric disorders (e.g., severe generalized anxiety disorder, bipolar disorder, schizophrenia etc.) where the condition is felt to be the most significant contributor to the cognitive impairment should be excluded. |
| 4. | Active medical conditions that impair cognition or impact participation in the study, other than AD should be exclusionary. Participants with medical conditions (e.g., severe obstructive sleep apnea) in whom the condition is felt to be the most significant contributor to the cognitive impairment should be excluded. |
| 5. | Significant neurodegenerative diseases, other than AD, and other causes of dementias. |
| 6. | A current diagnosis of uncontrolled type I or type II diabetes mellitus, as defined by Hemoglobin A1 C (Hb A1C > 8). |
| 7. | A current active, uncontrolled seizure disorder. |
| 8. | Participation in another clinical trial for an investigational agent of:<br>a. Symptomatic medications, participation is excluded within 4 weeks prior to the screening visit.<br>b. Disease-modifying medications, participation is excluded within 6 months of the screening visit. |
| 9. | Previous exposure to benfotiamine within past 3 months. |
| 10. | Current serious or unstable medical illness that could interfere with the interpretation of safety and assessment of efficacy in this study. |
| 11. | Diagnosis or treatment of cancer within the past 5 years. |
| 12. | History of alcoholism or substance abuse, current or within past 5 years. |
| 13. | Contraindication to MRI. |
| 14. | MRI at screening showing evidence of infection, tumor, cortical infarction, or multiple lacunes in prefrontal or critical memory regions. |
| 15. | Initiation of an FDA-approved monoclonal antibody treatment targeting brain amyloid (including lecanemab, aducanumab, solanezumab, donanemab) within 6 months prior to the screening visit. |
| 16. | A disability that may prevent the patient from completing all study requirements in the opinion of the PI. |

### Concomitant medications

Use of the following medications is prohibited throughout participation in the study: thiamine or benfotiamine supplements outside of a multivitamin, immunosuppressive medications, chemotherapeutic agents for malignancy, or any concomitant treatment which may impair cognitive function. Medications that are central nervous system active (e.g., hypnotics) and may affect cognitive function are not permitted for 72 hours prior to neuropsychological testing.

Eligible participants being treated with FDA-approved acetylcholinesterase inhibitors (AchEI) and/or memantine at the time of screening are expected to remain on a stable dosage regimen of these medications for the duration of the trial. They may be initiated as clinically indicated during the study when a participant is not already on these medications at the outset, with appropriate documentation. FDA-approved monoclonal antibodies targeting brain amyloid are permitted, but participants must be stable for at least 6 months on such treatment prior to screening. For medications not intended to treat AD, participants must be on a stable dose for at least 4 weeks prior to baseline, except for medications which are administered for a short course of treatment.

### Interventions

Benfotiamine capsules, 300 mg, 600 mg, or placebo are provided as opaque dark green (chlorophyll) colored capsules for oral administration. The placebo group will receive capsules with microcrystalline cellulose, with all other components, shape, and color identical to the active treatment. Participants are assigned capsules twice a day (BID; once in the morning and once in the evening). Participants receive a new supply for each dosing period between study visits and are provided a diary to track daily intake and timing of dose. During study visits, participant drug supply will be assessed for compliance.

### Randomization and blinding

A randomization schedule will be generated and incorporated into an electronic Randomization and Trial Supplies Management (RTSM) system. The treatment group will be assigned as the site randomizes the participant. In phase 2A of the trial, as shown in Figure 2, each participant will be randomly allocated in a 1:1:1 ratio into one of the 3 groups: 1200 mg/day benfotiamine, 600 mg/day benfotiamine or placebo. At the dose decision point that ends phase 2A, all participants enrolled in the two phase 2A active dose arms will receive a new supply of benfotiamine at the selected highest tolerable dose for phase 2B. An unscheduled visit at the start of phase 2B may be required to replace investigational product (IP) supply in all active participants once the phase 2A dose selection is made. Newly enrolled participants will be randomized 1:2 active arm to placebo arm until parity is achieved between numbers of active and placebo arm participants (approximately 150 participants total), randomized 1:1 thereafter (approximately 100 participants). In both phases of the trial, the randomization will be stratified by site.

Participants and study personnel will be blinded to both treatment arm and dosage. There will be provisions for the site Principal Investigator (PI) to unblind treatment assignment in consultation with the ADCS Medical Monitor only in the case of an emergency.

### Adverse events

All AEs (i.e., a new event or an exacerbation of a pre-existing condition in its frequency or severity) that occur from the time of consent and up to 30 days after the IP has been discontinued will be recorded. A treatment emergent AE (TEAE) is defined as any AE that developed, worsened, or became serious after first dose of IP and prior to 30 days after the last dose of IP. A serious adverse event (SAE) is an AE that results in any of the following outcomes: death, life-threatening situation (participant is at immediate risk of death), inpatient hospitalization or prolongation of existing hospitalization, persistent or significant disability/incapacity, congenital anomaly/birth defect in the offspring of a patient who received IP, or considered significant by the investigator for any other reason. All AEs will be followed until resolution or until the Investigator and/or the Medical Monitor determine the event is chronic or clinically stable.

### Safety Surveillance

During both phases of the trial, safety surveillance includes monitoring of 1) AEs/TEAEs/SAEs summarized by grade, attribution, and body systems, by number of events, and by participant, 2) physical exam abnormalities, 3) vital signs (weight, blood pressure, pulse rate, temperature, respiration rate), 4) standard 12-lead resting ECG measures, and 4) summaries of safety laboratory tests.

### Outcomes

#### Phase 2A primary endpoints

The primary safety endpoint in phase 2A is the rate of TEs in active and placebo arms at each dose. A TE is defined as early discontinuation of the IP due to intolerability or a post-randomization moderate or severe AE that is possibly, probably or definitely related to the IP.

#### Phase 2A secondary endpoints

Secondary safety and tolerability endpoints for phase 2A include: 1) number of participants with AEs/TEAEs and SAEs, 2) number of participant withdrawals from the study, and 3) number of participants discontinuing IP.

#### Phase 2B primary endpoints

The primary cognitive endpoint is the within-participant change from baseline to 72 weeks compared between active arm and placebo on the ADAS-Cog13 [23, 24]. The primary functional endpoint is the within-participant change from baseline to 72 weeks compared between active arm and placebo on the CDR-Sum of Boxes (SB) [25, 26].

#### Phase 2B secondary safety and tolerability endpoints

Secondary safety and tolerability endpoints for phase 2B include: 1) number of participants with AEs/TEAEs and SAEs 2) number of participant withdrawals from the study, and 3) number of participants discontinuing IP.

#### Phase 2B secondary PK endpoints

Secondary PK endpoints for phase 2B include mean and median levels of: 1) thiamine (nmol/L), 2) thiamine diphosphate (ThDP) (nmol/L), 3) thiamine monophosphate (ThMP) (nmol/L) [27], and 4) erythrocyte transketolase activity (U/g haemoglobin (U/gHb)) [28, 29]. Measures of ThDP, ThMP, and erythrocyte transketolase activity will be provided as blood markers of efficacy of drug delivery. These measurements will be conducted on whole and red blood cells in samples collected at baseline, week 8, and week 72.

#### Phase 2B secondary efficacy endpoints

Additional efficacy endpoints of benfotiamine will be assessed by within-participant change from baseline to week 72 between active arm and placebo on the following: 1) the Alzheimer’s Disease Cooperative Study – Activities of Daily Living Scale for use in Mild Cognitive Impairment (ADCS-ADL-MCI) [30] to evaluate preservation or potential improvement in everyday activities of daily living, and 2) the MoCA [31] to evaluate preservation or potential improvement in cognitive function.

#### Phase 2B exploratory endpoints

Exploratory endpoints in phase 2B include within-participant longitudinal change between active arm and placebo in: 1) cranial MRI volumetric measures of hippocampal, whole brain, and ventricles, as well as regional cortical thickness, 2) scores on the Neuropsychiatric Inventory (NPI) [32], 3) scores on neuropsychological test measures administered remotely and in-person, 4) ADAS-Cog-Exec [33] scores, 5) levels of plasma biomarkers Aβ42/Aβ40 ratio, p- tau217/np-tau217 ratio, p-tau217, total tau, p-tau 231, NfL and GFAP, 6) AGE levels measured in plasma as an exploratory endpoint of PD, and 7) PK (plasma thiamine, thiamine esters in whole blood including ThDP and ThMP, and erythrocyte transketolase activity) relationships to PD biomarkers (AGE) and clinical outcome measures.

AGE levels will be measured on plasma samples and four AGE by mass spectrometry (N(6)-carboxymethyl-lysine (CML), pentosidine, N(6)-carboxymethyl-lysine (CEL), methylglyoxal-derived hydroimidazolone) will be quantified to measure all specific modes of action of benfotiamine.

Neuropsychological measures administered in-person include the Boston Naming Test (30-item) [34], the Trail Making Test (Parts A and B) [35], and WAIS-R Digit Symbol Substitution [36]. Neuropsychological measures administered remotely (via video-call) include Immediate and Delayed Recall from the Rey Auditory Verbal Learning Test [37], Number Span Forward and Backward [38], Category Fluency [39] and Letter Fluency [39]. Remote measures will be administered over videoconference-call within 10 days after each in-person visit (baseline, week 24, week 48, and week 72). Participants will use their own interactive video capable devices for the remote assessments, although study-provided devices will be available, as needed.

### Power calculation and sample size

Table 3 shows that with a total sample size of n=406 (203/arm), a 20% dropout rate, and a 5% drop-in rate from the placebo arm to active treatment, the trial will be powered at 80% overall (90% on each endpoint separately) to detect a difference between arms, corresponding to a Cohen’s d effect size of 0.38 which represents a small to moderate effect size. This would be a reasonable and clinically relevant difference between arms on each co-primary endpoint, and would be commensurate to the 35% to 40% amelioration of decline seen in the placebo arm of the recently completed phase 2 trial of donanemab in patients with early AD [40], and would correspond to an absolute difference between arms on the CDR-SB of 0.7 points (assumed SD = 1.74) and of 1.7 points (assumed SD = 4.35) on the ADAS-Cog13.

**Table 3.** Sample sizes which achieve 90% power for given effect sizes (Cohen’s D; difference between the 2 arms), assuming 20% dropout and assuming drop-in rates of 5% and 10%.

| drop-in rate | effect size | n / arm |
| --- | --- | --- |
| 5% | 0.35 | 239 |
| **5%** | **0.38** | **203** |
| 5% | 0.40 | 183 |
| 10% | 0.35 | 266 |
| 10% | 0.38 | 226 |
| 10% | 0.40 | 204 |

### Statistical analyses

#### Phase 2A

When criteria have been reached to end phase 2A, the unblinded safety biostatistician will alert the PIs (HHF, JAL, GEG) that the database is to be frozen. An unblinded safety report will be provided to the Data Safety Monitoring Board (DSMB). The phase 2A primary analysis will compare the rate of TEs between the 1200 mg/day arm and the placebo arm. The analysis will test whether the rate of TEs is unacceptably high in this high-dose arm compared to placebo, using a formal one-sided hypothesis test of equal rates, against the one-sided alternative that the high dose arm TE rate is greater than the placebo arm TE rate. The conditional one-sided test of Przyborowski and Wilenski [41] will be used, at 5% significance level.

Additional phase 2A safety analyses will involve computing an exact upper-one sided 95% confidence interval for the TE rate in each arm. Safety and tolerability data and demographic data will be summarized in a tabular and/or graphical format for each treatment group. The incidence of all laboratory test abnormalities and median changes from baseline will be tabulated by treatment regimen and time point. The safety population (i.e., enrolled and randomized participants who received at least 1 dose of blinded study therapy) will be used for these analyses.

#### Phase 2B

The primary analyses will be performed on the modified intent to treat (mITT) population which includes all randomized participants who took at least one dose of the study medication, have a baseline assessment of the co-primary efficacy endpoints, and have at least one efficacy evaluation following baseline. The primary analysis will be the same for each co-primary endpoint (CDR-SB and ADAS-Cog13) and will test within subject change in the measure as the outcome in a mixed effects repeated measures model using all available outcomes in the mITT population with no imputation for missing data. Fixed effects in the model will be *APOE* status, baseline value of the outcome measure, baseline by visit-week interaction term, treatment group, visit-week, and visit-week by treatment group interaction. The approach will be maximum likelihood. Visit will be treated as a categorical variable. The covariance structure will be specified as follows: the random effects will include site and subject. The within-subject covariance will be unstructured. If needed, sites will be pooled (in order of enrollment, starting from minimum enrollment) so that there is a minimum of 5 participants per site. If the model fails to converge at the default setting for the software used, then site will be changed to a fixed effect, and if this model does not converge, site will be removed from the model. If the model does not converge, the following structures for the within subject covariance will be fit, sequentially, until the structure is found that results in convergence of the model: Huynh-Feldt, Toeplitz, Autoregressive (1), and Compound Symmetry. A sandwich estimator will be utilized to estimate the variance estimates, and degrees of freedom will be calculated using the between-within method. The primary endpoint will be tested using model-adjusted least squares means at the week 72 visit. Point estimates, standard errors, two-sided 95% confidence intervals, and p-values will be presented.

Mean changes in the secondary endpoints from baseline to week 72 will be compared between arms using the analysis strategy outlined above. To control alpha over the primary and all secondary endpoints, an overall hierarchical gatekeeper strategy will be used, similar to the primary analysis. First, if the primary analysis is not significant, then the secondary endpoints will not be formally tested, and will be presented using exploratory summary statistics only (means and uncorrected 95% confidence intervals by arm and for the difference between arms). No claims of statistical significance will be made. However, if the primary analysis is significant, the secondary endpoints will be tested in the listed order in hierarchical manner, each using an alpha of 5%. Once a secondary endpoint has failed to attain the prespecified 5% significance level, the remaining secondary endpoints will be presented as exploratory only. This strategy will maintain the overall familywise significance level for the primary and all secondary endpoints at 5%. Exploratory analyses will be detailed in the statistical analysis plan, and will generally be similar to the primary and secondary analyses.

Safety and tolerability data as well as demographic data for the safety population will be summarized in tabular and/or graphical format for each treatment group. The incidence of all laboratory test abnormalities and the median changes from baseline will be tabulated by treatment regimen and time point.

### Data safety monitoring board

The DSMB will provide safety oversight of the trial. It is independent from the ADCS and reports to the National Institute on Aging (NIA) through its project scientist. The DSMB includes a chair and 4 other experts in the field who have the appropriate background for these responsibilities and have been approved by the NIA. Other members may be added depending on the needs of the trial. The DSMB will meet quarterly and on an ad-hoc basis in the face of any important safety matters that may arise, and will provide recommendations following each meeting. The final recommendation on dose to select at the end of phase 2A for use in phase 2B will be made by the DSMB. The DSMB will have the authority to recommend that the study be halted or discontinued.

### Early discontinuation

The entire study may be otherwise discontinued at the discretion of the ADCS or the study PIs (GEG, JAL, HHF) for administrative or other reasons. Participants are free to withdraw from study participation at any time, for any reason, and without prejudice. Discontinuation of study treatment and/or the participation of an individual patient can occur in the following circumstances: 1) withdrawal of informed consent by the participant or legally authorized representative (LAR), 2) treatment with a non-permitted concomitant medication, 3) AE or other significant medical condition which, in the opinion of the Investigator, render it necessary to discontinue study drug, 4) medical emergency that necessitates unblinding the participant’s treatment assignment, 5) any other occurrence that, in the PIs opinion, makes continued participation contrary to the participant’s best interest, or 6) movement of the participant into a long-term care nursing facility. Participants who discontinue study treatment for any reason will have the opportunity to continue on the protocol to the end of the study with their ongoing consent.

### Monitoring & quality control

During the study each site will be monitored at regular intervals by an ADCS clinical research monitor through a combination of on-site visits and remote monitoring procedures. The monitoring visits will be conducted according to the applicable International Conference on Harmonization (ICH) and Good Clinical Practice (GCP) guidelines to ensure protocol adherence, quality of data, drug accountability, compliance with regulatory requirements, and continued adequacy of the investigation site and its facilities. Documentation of all quality control procedures will be outlined in the Data Management Plan. Edit checks and listings will be run and used in conjunction with electronic case report form pages to support clinical review of the data.

### Ethics

Review and approval of the protocol and the associated informed consent documents and recruitment materials was conducted by the central Institutional Review Board (IRB) Advarra. Approval from the central IRB will be obtained for all protocol amendments and changes to the informed consent document when they are warranted. The site investigator will notify the central IRB of deviations from the protocol or SAEs occurring at the site.

No study-specific procedure will be undertaken on any individual patient until that patient or their LAR has given written informed consent to take part in the study. The study partner must also participate in the consenting process. Participant confidentiality is strictly held in trust by the participating investigators, research staff, and the sponsoring institution and their agents.

### Trial Status

Recruitment for this study began March 22, 2024 and is currently ongoing.

### Dissemination plan

Study results will be published in accordance with the 2010 Consolidated Standards of Reporting Trials (CONSORT) guidelines. As there are expected to be too few participants at each site for an individual site’s results to be statistically valid, the results of this study will be disclosed or published only in combined form based upon the statistical analysis coordinated and performed by the ADCS. Results will be posted on clinicaltrials.gov in accordance with requirements.

## Discussion

There is an urgent need to accelerate and diversify therapeutic possibilities for patients with AD [8]. Benfotiamine provides an important novel therapeutic direction in AD as it addresses and treats a well-characterized tissue thiamine action deficiency and related changes in glucose metabolism as well as post-translational modifications including those promoting plaque [2] and tangle formation [3], neuroinflammation [4, 5], neurodegeneration [6], and AGE [7]. Benfotiamine has potential to achieve POC, with the possibility of additive or synergistic effects to other current therapeutic approaches, addressing the US National Alzheimer’s Project Act (NAPA) goal of combining targets [8]. The preliminary results of a pilot study provide proof of principle that justify testing the efficacy of benfotiamine in a larger RCT to investigate its safety, tolerability, and efficacy in early AD [6, 18].

The BenfoTeam trial is deploying an innovative seamless phase 2A-2B design to achieve POC. This is a double-blind, placebo-controlled RCT of 72-weeks of benfotiamine in early AD with a randomized sample of 406 participants. It includes an adaptive dose decision rule that optimizes exposure to the highest and best-tolerated dose. An additional innovation is the use of plasma-based amyloid and p-tau biomarker tests for inclusion, which provides an important contribution by shifting away from invasive testing using lumbar puncture or expensive PET scanning that entails significant radiation exposure. This phase 2A-2B trial is powered to provide evidence of clinically important benefit and, if positive, it will further validate our approach of deploying seamless designs with adaptive dose finding, and will provide measures and effect sizes that can become new benchmarks.

Results from the BenfoTeam trial are expected to identify the highest well-tolerated dose of benfotiamine that has an acceptable safety profile. Measurable PK and PD biomarkers and a constellation of downstream plasma AD and neurodegenerative biomarkers ensure that the trial will be able to test the sufficiency of target engagement and dose while evaluating longer term efficacy and safety. In addition, the sample size is well powered to determine if benfotiamine will benefit cognition and everyday function in individuals with early AD.

## Data Availability

This protocol does not report experimental results and data collection is ongoing. De-identified data will be made available to qualified researchers upon study completion, following publication of the primary manuscript. URL: https://www.adcs.org/data-sharing/

## Authors Contributions

**Conceptualization:** Howard H. Feldman, Gary E. Gibson, José A. Luchsinger.

**Project Directors/Principal Investigators:** Howard H. Feldman, Gary E. Gibson, José A. Luchsinger.

**Funding Acquisition:** Howard H. Feldman, Gary E. Gibson [contact principal investigator], José A. Luchsinger.

**Methodology:** Howard H. Feldman, Gary E. Gibson, José A. Luchsinger, Karen Messer, Steven D. Edland, Diane M. Jacobs, David P. Salmon, Sarah A. Flowers, Kerry S. Jones, Albert Koulman, Philip B. Verghese, Venky Venkatesh, Kevin Yarasheski, Henrik Zetterberg.

**Project Administration:** Carolyn Revta.

**Writing – Original Draft Preparation:** Howard H. Feldman, January Durant, Jody-Lynn Lupo.

**Writing – Review & Editing:** Howard H. Feldman, Gary E. Gibson, José A. Luchsinger, Gabriel C. Léger, Curtis Taylor, Diane M. Jacobs, David P. Salmon, Karen Messer, Steven D. Edland, Carolyn Revta, Sarah A. Flowers, Kerry S. Jones, Albert Koulman, Philip B. Verghese, Venky Venkatesh, Kevin Yarasheski, Henrik Zetterberg, January Durant, Jody-Lynn Lupo.

## Acknowledgments

The members of the ADCS BenfoTeam Study Group are as follows:

**ADCS:** Department of Neurosciences, Alzheimer’s Disease Cooperative Study, University of California, San Diego, CA, USA.

**ADCS Co-Directors:** Howard H. Feldman, Judy Pa.

**Project Directors:** Gary E. Gibson (contact PI), José A. Luchsinger, Howard H. Feldman.

**Executive Operations:** Carolyn Revta.

**Clinical Operations:** Savannah Aries, Briana Askew*, Kemi Badru*, Veronica Fernandez, Nisha Johal*, Tamesha Lucas, Bryce Truver, Erika Velazco.

**Clinical Monitoring:** R. Michelle Brill, Laura Cole, Edward Fox, Ben Germain, Don Guterwill, Bailey Hoffman, Tiffany Jones, Janet Kastelan, Ike Monye*, Susan Mroz, Gerald Rivera*, Rebecca Ryan-Jones*, Vicky Syed*, Kheshini De Zoysa.

**Regulatory/Quality Management:** Pat Chinwattana, Lori Houghtaling, Roxana Phillips, Teresa Ruiz, Rick Seghers.

**Data Management:** Susan Castellino*, Sneha Jasti, Jim Krooskos, Andrew MacKelfresh, Keerthi Vemuri, Zel Zorin.

**Medical Safety:** Gabriel Leger, Alessandra Pol, Curtis Taylor.

**Instruments:** Diane M. Jacobs, David P. Salmon.

**Recruitment:** Daniel Bennett, Andrea LaCroix, Genevieve Matthews, Donna Tan.

**Imaging:** James Brewer, Nichol Ferng, Robin Jennings.

**Statistics:** Steven D. Edland, Shelia Jin, Karen Messer, Jing Zhang.

**Special Projects/Medical Writing:** January Durant, Jody-Lynn Lupo.

**Biomarker/Lab Studies:** Sarah A. Flowers, University of Virginia, Charlottesville, Virginia, United States of America; Kerry S. Jones, Damon A. Parkington, Sarah R. Meadows and Albert Koulman, University of Cambridge, Cambridge, United Kingdom; Kevin Yarasheski, Philip B. Verghese, and Venky Venkatesh, C_2_N Diagnostics, St. Louis, Missouri, United States of America; Henrik Zetterberg, Sahlgrenska Academy at the University of Gothenburg, Mölndal, Sweden.

*no longer active at ADCS or with this project

## Supporting Information

S1 File. **Statistical Methods: Transition from Phase 2A to Phase 2B**

S2 File. **Data sharing statement**

S3 File. **The protocol approved by the ethics committee**

S4 File. **Model consent form**

S1 Checklist. **SPIRIT 2013 checklist**

